# Quantifying changes in the IFR and IHR over 23 months of the SARS-CoV-2 pandemic in England

**DOI:** 10.1101/2022.10.12.22281016

**Authors:** Oliver Eales, David Haw, Haowei Wang, Christina Atchison, Deborah Ashby, Graham Cooke, Wendy Barclay, Helen Ward, Ara Darzi, Christl A. Donnelly, Marc Chadeau-Hyam, Paul Elliott, Steven Riley

## Abstract

**Background:** The relationship between prevalence of infection and severe outcomes such as hospitalisation and death changed over the course of the COVID-19 pandemic. The REal-time Assessment of Community Transmission-1 (REACT-1) study estimated swab positivity in England approximately monthly from May 2020 to 31 March 2022. This period covers widespread circulation of the original strain, the emergence of the Alpha, Delta and Omicron variants and the rollout of England’s mass vaccination campaign.

**Methods:** Here, we explore this changing relationship between prevalence of swab positivity and the infection fatality rate (IFR) and infection hospitalisation rate (IHR) over 23 months of the pandemic in England, using publicly available data for the daily number of deaths and hospitalisations, REACT-1 swab positivity data, time-delay models and Bayesian P-spline models. We analyse data for all age groups together, as well as in two sub-groups: those aged 65 and over and those aged 64 and under.

**Results:** During 2020, we estimated the IFR to be 0.67% and the IHR to be 2.6%. By late-2021/early-2022 the IFR and IHR had both decreased to 0.097% and 0.76% respectively. Continuous estimates of the IFR and IHR of the virus were observed to increase during the periods of Alpha and Delta’s emergence. During periods of vaccination rollout, and the emergence of the Omicron variant, the IFR and IHR of the virus decreased. During 2020, we estimated a time-lag of 19 days between hospitalisation and swab positivity, and 26 days between deaths and swab positivity. By late-2021/early-2022 these time-lags had decreased to 7 days for hospitalisations, and 18 days for deaths.

**Conclusion:** Even though many populations have high levels of immunity to SARS-CoV-2 from vaccination and natural infection, waning of immunity and variant emergence will continue to be an upwards pressure on IHR and IFR. As investments in community surveillance are scaled back, alternative methods should be developed to accurately track the ever changing relationship between infection, hospitalisation and death.

## Introduction

Since its first detection in late 2019, SARS-CoV-2 has led to high levels of morbidity and mortality worldwide [1,2]. In England in late 2020, following the emergence of the Alpha variant, which has been linked to higher levels of transmissibility [3,4] and severity [5], there was a rapid rise in infections leading to a surge in hospitalisations and deaths [6] and to intense pressure on the national health service (NHS). In order to stem the tide of infections a national lockdown was introduced on 6 January 2021 [7] aimed at drastically reducing social contacts. Simultaneously, as lockdown began, England began implementing a mass vaccination campaign and has since reached high levels of vaccine coverage [6]. Studies have found the vaccines to be highly effective against severe outcomes at the individual level [8] and have also been linked to reduced levels of transmission [9]. The combined effect of both lockdown and vaccinations led to a sharp decrease in cases, hospitalisations, and deaths during the first few months of 2021 [6].

After March 2021, lockdown restrictions were slowly lifted with phased reopenings [10]. Combined with the emergence of the Delta variant in England in April 2021, which has been linked to even greater levels of transmissibility [11,12] and reduced vaccine effectiveness [13], the pandemic once more entered a phase of growth leading to higher levels of prevalence [14]. The last easing on 19 July 2021 removed all legal restrictions and saw society reopen to an extent not seen since March 2020 [15]. Restrictions have not since been implemented at such a large scale in England, despite high continuous levels of prevalence during the summer and autumn of 2021 [16], and two large waves of infection [17] following the emergence of the Omicron variant and its BA.1 and BA.2 sub-lineages respectively [16]. In deciding to implement or remove restrictions, the UK government made their main criteria that “Infection rates do not risk a surge in hospitalisations which would put unsustainable pressure on the NHS” [10]. Assessing trends between levels of infection and hospitalisation rates is therefore crucial in better informing governments and public health bodies so that restrictions can be appropriate and proportionate.

The infection fatality ratio (IFR) and infection hospitalisation ratio (IHR) measure the proportion of deaths and hospitalisations respectively among infected individuals. When they are known accurately, short-term forecasts of severe outcomes can be made using current estimates of infection levels and the estimated time-delay to severe outcomes [18,19].

Models forecasting future levels of infection [20–22] can also then easily be converted into forecasts of severe outcomes. Such forecasts can allow health care services to better prepare for periods of increased capacity, through better scheduling and procurement of sufficient levels of equipment. Accurate estimates of the IFR and IHR have been made during the pandemic [23,24]. However, with the introduction of new variants, the rollout of vaccinations, the waning of vaccine effectiveness [25–27], and the effect of vaccine booster doses [28] the values of IFR and IHR can rapidly change without necessarily being easily detected.

The REal-time Assessment of Community Transmission-1 (REACT-1) study is a repeat cross-sectional survey that aimed to test a random sample of the England population for the presence of the SARS-CoV-2 virus at fixed points in time [29]. A round of the study occurred approximately monthly between May 2020 and March 2022 with between 95,000 and 175,000 participants each round. The study allowed the progression of the pandemic in England to be accurately measured [30] without the biases of normal sampling methods [31].

By accurately characterising the relationship between severe outcomes and this gold-standard data, real-time changes in the severity of the virus can be detected and quantified with only a small delay (due to the time between infection and a severe outcome). We present here the inferred relationship between the prevalence estimated from round 1 to round 19 of REACT-1 (May 2020 – April 2022), and the daily number of deaths and hospital admissions in England over the same period, using a simplistic time-delay model.

## Results

### Quantifying the relationship between swab positivity and severe outcomes

The time-delay between swab positivity and severe outcomes decreased over the duration of the study. Fitting the time-delay model (see Methods) to rounds 1-7 (1 May - 3 December 2020) of REACT-1 we estimated a discrete time-lag of 19 (18, 20) days to hospitalisations, and a time-lag of 26 (25, 27) to deaths (Figure 1, Supplementary Table 1). These estimates were in line with those obtained by fitting the same model to all 19 rounds of the data. Fitting the model to rounds 14-19 (9 September 2021 - 1 April 2022) of the study, we estimated much shorter time-lags of 7 (7, 8) days to hospitalisations and 18 (18, 18) days to deaths.

**Figure 1.**
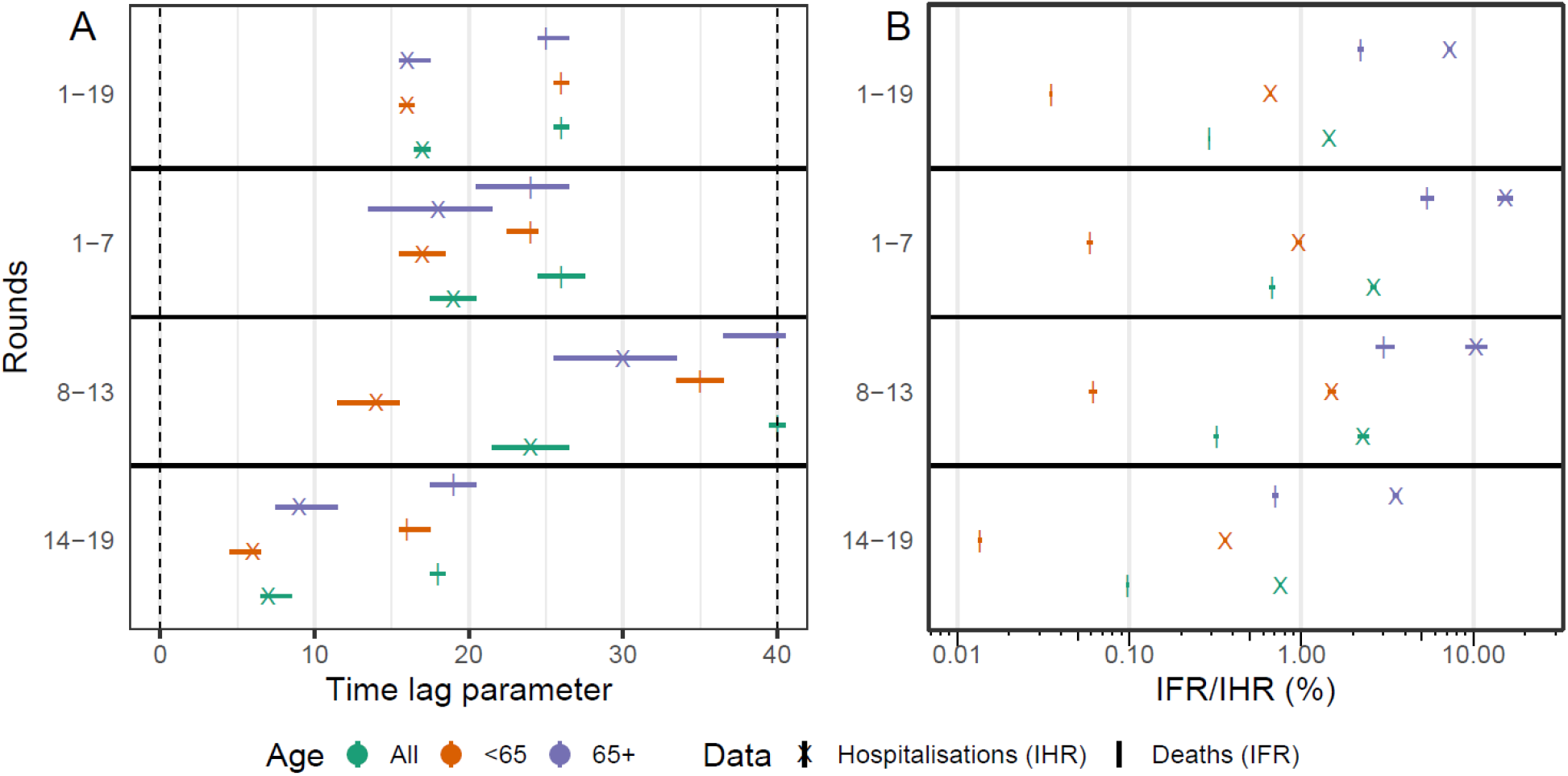
Comparison of parameters obtained by fitting to different subsets of the REACT-1 data. **(A)** Best-fitting time-lag parameter and 95% credible intervals for models fit to deaths (cross) and hospitalisations (vertical line), for models fit to all age-groups (green), those aged 64 and under (orange) and those aged 65 and over (purple). **(B)** Best-fitting scaling parameter and 95% credible intervals on a *log*_10_x-axis for models fit to deaths (cross) and hospitalisations (vertical line), for models fit to all age-groups (green), those aged 64 and under (orange) and those aged 65 and over (purple).

Models fit to subsets of the data by age group (64 years and under, 65 years and over) showed similar time-lags between age-groups, though the time-lags tended to be slightly shorter for those aged 64 and under. In contrast, fitting the models to round 8-13 (30 December 2020 - 12 July 2021) of REACT-1 identified wildly different estimates between age groups, and some extremely long time-lags. However, this was over a period of time when large proportions of the population were being vaccinated against COVID-19, with different rates of vaccination by age group (Figure 2). This could have led to continuously changing severity, massively biassing estimates obtained from the model. Due to the likely high degree of bias during rounds 8-13, below we present only the models obtained from rounds 1-7 and rounds 14-19.

**Figure 2.**
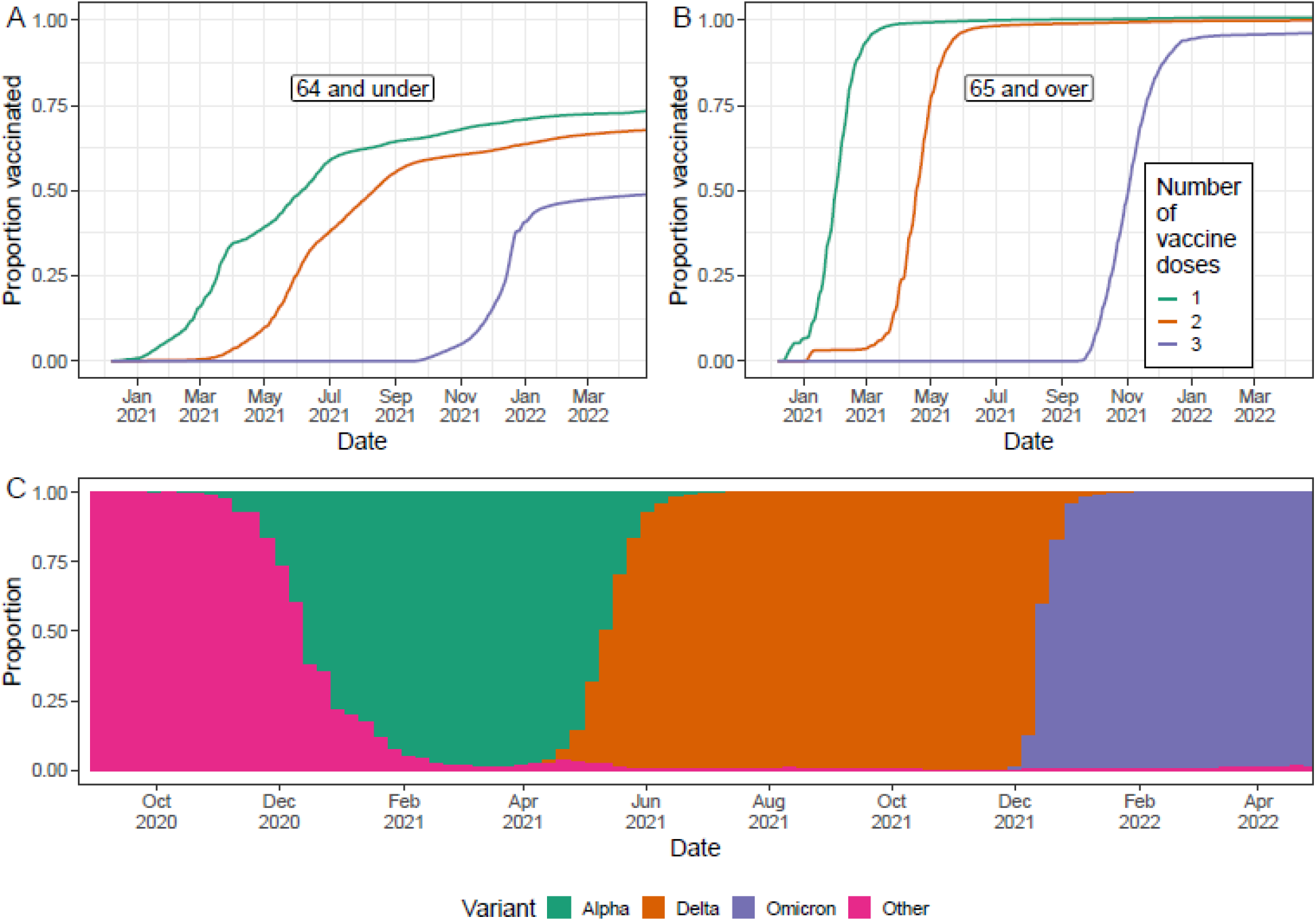
Vaccination coverage and variants responsible for infections in England, as inferred from public data sets. **(A, B)** Proportion of individuals who have been vaccinated with at least a single dose (green), proportion of individuals who have been vaccinated with at least two doses (orange), and the proportion of individuals who have received a third ‘booster’ dose (purple), for those aged 64 and under (A) and those aged 65 and over (B). **(C)** Proportion of infections in England, for which lineages were detected, that were identified as the Omicron variant (purple), the Delta variant (orange), the Alpha variant (green) and any other lineage (pink).

There was a high degree of correlation between smoothed estimates of swab positivity and the time-delayed signal of severe outcomes, though over the course of the study the two signals substantially diverged (Figures 3-5, Supplementary Table 2). Assuming the estimated time-lag for the time-delay models fit to rounds 1-7 of REACT-1, we found a Pearson correlation for modelled swab positivity over rounds 1-7 against death and hospitalisations of 0.985 and 0.978 respectively (p-values<0.001). Similarly high levels of correlation were found for rounds 8-13 assuming the same time-lag. Correlation over rounds 14-19 was moderately lower when assuming a time-lag from models fit to rounds 1-7 (though still significant, p-values<0.001), but only slightly lower when assuming the time-lags from models fit to rounds 14-19. Broadly similar patterns in correlations were found when looking at both different age-groups. However, for those aged 64 and under during rounds 14-19 correlation was greatly reduced with a value of 0.169 (p-value=0.01), even when assuming the time-lag from the model fit to rounds 14-19.

**Figure 3.**
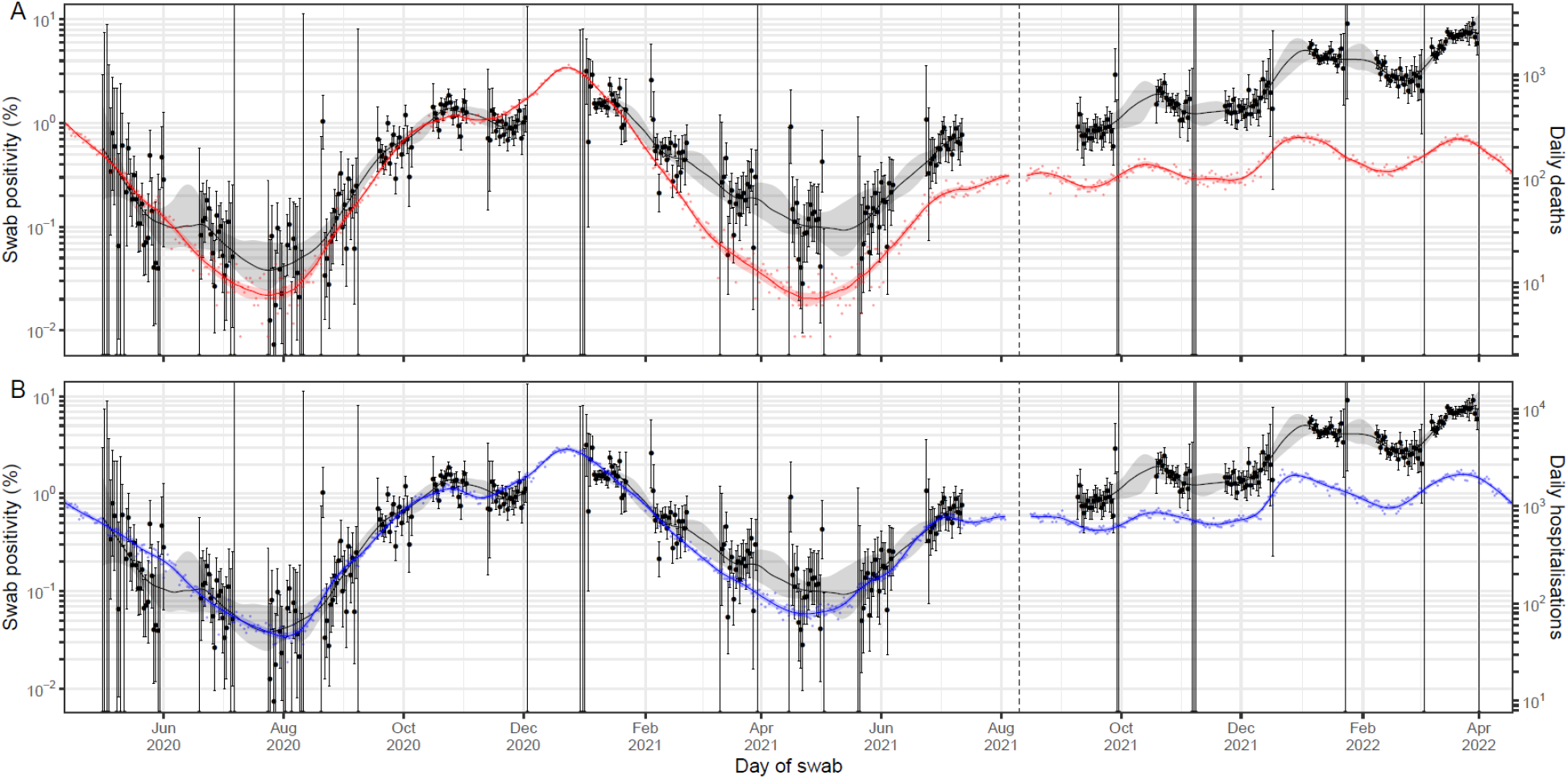
A comparison of daily deaths and hospitalisations to swab positivity as measured by REACT-1. Daily swab positivity for all 19 rounds of the REACT-1 study (black points with 95% credible intervals, left hand y-axis) with P-spline estimates for swab positivity (solid black line, shaded area is 95% credible interval). **(A)** Daily deaths in England (red points, right hand y-axis) and P-spline model estimates for expected daily deaths in England (solid red line, shaded area is 95% credible interval, right hand y-axis). The black vertical dashed line on 10 August 2021 splits the data into two periods: rounds 1-13 and rounds 14-19 of REACT-1. During rounds 1-13 daily deaths have been shifted by 26 days backwards in time along the x-axis. During rounds 14-19 daily deaths have been shifted by 18 days backwards in time along the x-axis.The two y-axis have been scaled using the population size and best-fit scaling parameter from the time-delay model fit to rounds 1-7. **(B)** Daily hospitalisations in England (blue points, right hand y-axis) and P-spline model estimates for expected daily hospitalisations in England (solid blue line, shaded area is 95% credible interval, right hand y-axis). The black vertical dashed line on 10 August 2021 splits the data into two periods: rounds 1-13 and rounds 14-19 of REACT-1. During rounds 1-13 daily hospitalisations have been shifted by 19 days backwards in time along the x-axis. During rounds 14-19 daily hospitalisations have been shifted by 7 days backwards in time along the x-axis. The two y-axis have been scaled using the population size and best-fit scaling parameter from the time-delay model fit to rounds 1-7 of REACT-1.

**Figure 4.**
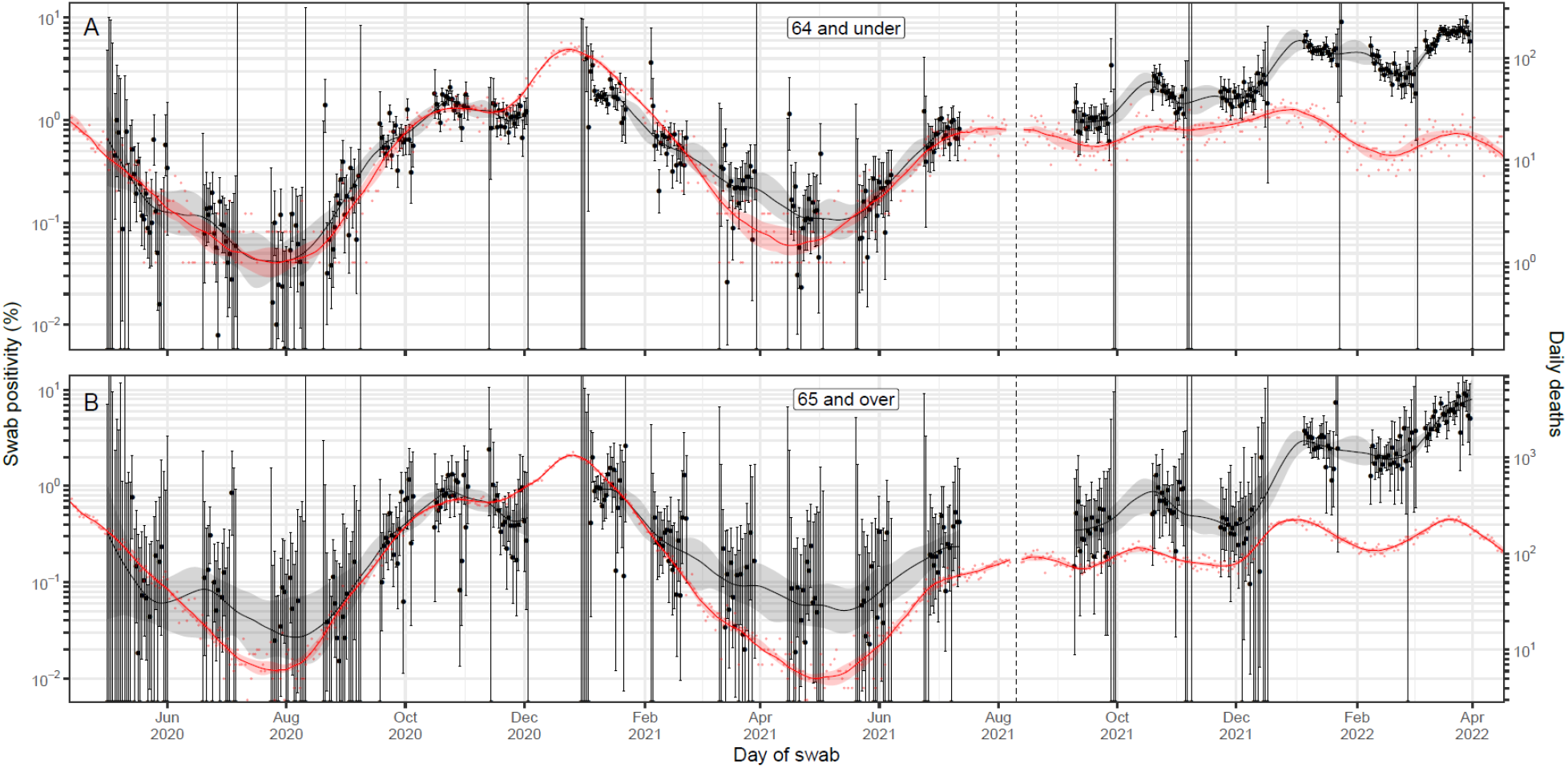
A comparison of daily deaths to swab positivity as measured by REACT-1, by age group. Daily swab positivity for all 19 rounds of the REACT-1 study (black points with 95% credible intervals, left hand y-axis) with P-spline estimates for swab positivity (solid black line, shaded area is 95% credible interval) for (A) those aged 64 and under, and (B) those aged 65 and over. **(A)** Daily deaths for those aged 64 and under in England (red points, right hand y-axis) and corresponding P-spline model estimates for the expected number of deaths (solid red line, shaded area is 95% credible interval, right hand y-axis). The black vertical dashed line on 10 August 2021 splits the data into two periods: rounds 1-13 and rounds 14-19 of REACT-1. During rounds 1-13 daily deaths have been shifted by 24 days backwards in time along the x-axis. During rounds 14-19 daily deaths have been shifted by 16 days backwards in time along the x-axis.The two y-axis have been scaled using the population size and best-fit scaling parameter from the time-delay model fit to rounds 1-7 of REACT-1. **(B)** Daily deaths for those aged 65 and over in England (red points, right hand y-axis) and corresponding P-spline model estimates for the expected number of deaths (solid red line, shaded area is 95% credible interval, right hand y-axis). The black vertical dashed line on 10 August 2021 splits the data into two periods: rounds 1-13 and rounds 14-19 of REACT-1. During rounds 1-13 daily deaths have been shifted by 24 days backwards in time along the x-axis. During rounds 14-19 daily deaths have been shifted by 19 days backwards in time along the x-axis.The two y-axis have been scaled using the population size and best-fit scaling parameter from the time-delay model fit to rounds 1-7 of REACT-1.

**Figure 5.**
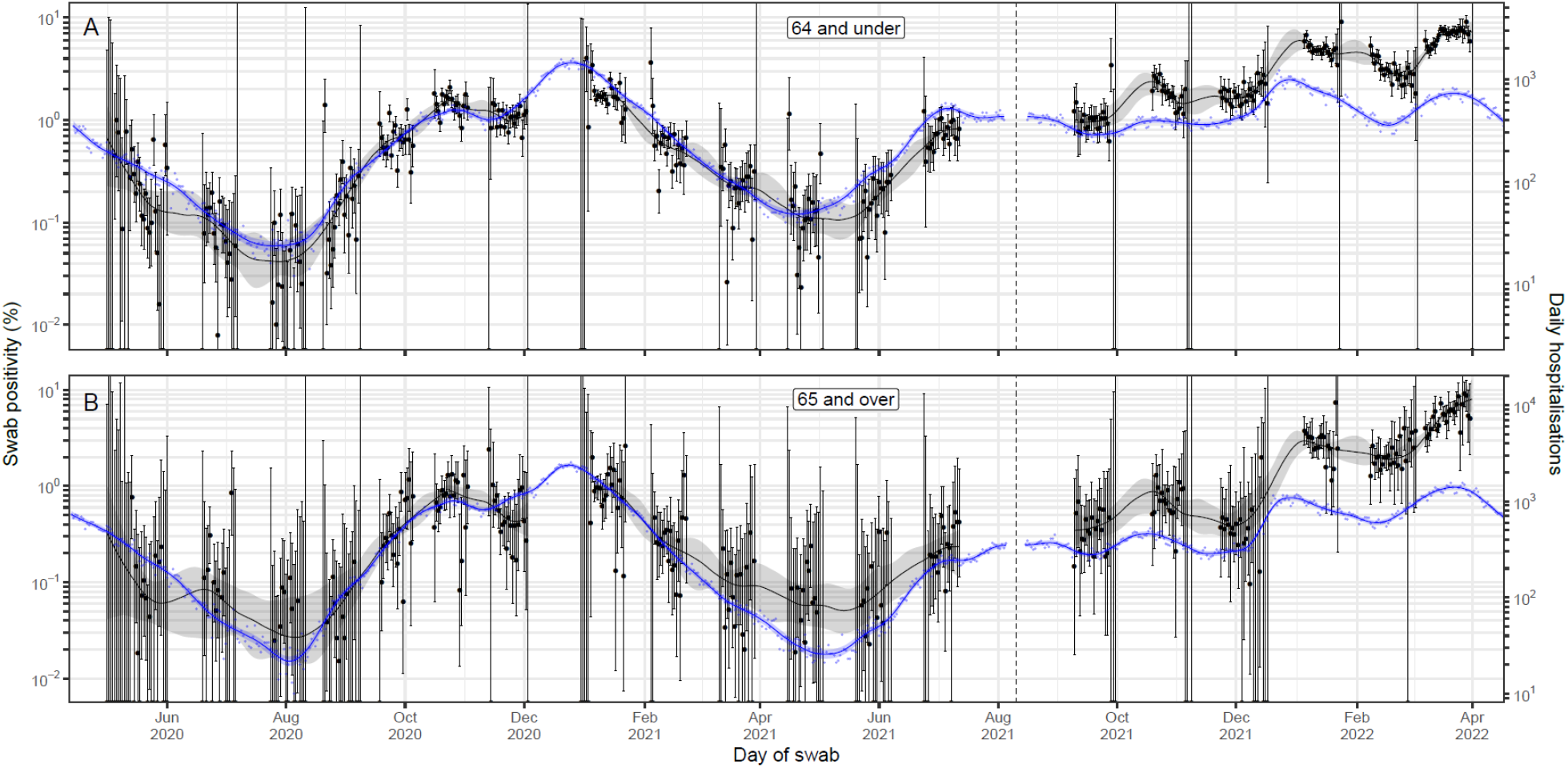
A comparison of daily deaths to swab positivity as measured by REACT-1, by age group. Daily swab positivity for all 19 rounds of the REACT-1 study (black points with 95% credible intervals, left hand y-axis) with P-spline estimates for swab positivity (solid black line, shaded area is 95% credible interval) for (A) those aged 64 and under, and (B) those aged 65 and over. **(A)** Daily hospitalisations for those aged 64 and under in England (blue points, right hand y-axis) and corresponding P-spline model estimates for the expected number of hospitalisations (solid blue line, shaded area is 95% credible interval, right hand y-axis). The black vertical dashed line on 10 August 2021 splits the data into two periods: rounds 1-13 and rounds 14-19 of REACT-1. During rounds 1-13 daily hospitalisations have been shifted by 17 days backwards in time along the x-axis. During rounds 14-19 daily hospitalisations have been shifted by 6 days backwards in time along the x-axis. The two y-axis have been scaled using the population size and best-fit scaling parameter from the time-delay model fit to rounds 1-7 of REACT-1. **(B)** Daily hospitalisations for those aged 65 and over in England (blue points, right hand y-axis) and corresponding P-spline model estimates for the expected number of hospitalisations (solid blue line, shaded area is 95% credible interval, right hand y-axis). Daily hospitalisations have been shifted by 18 days backwards in time along the x-axis. The black vertical dashed line on 10 August 2021 splits the data into two periods: rounds 1-13 and rounds 14-19 of REACT-1. During rounds 1-13 daily hospitalisations have been shifted by 18 days backwards in time along the x-axis. During rounds 14-19 daily hospitalisations have been shifted by 9 days backwards in time along the x-axis. The two y-axis have been scaled using the population size and best-fit scaling parameter from the time-delay model fit to rounds 1-7 of REACT-1.

The estimated severity of the virus was found to decrease over the duration of the study. Using the time-delay model we were able to estimate the IFR and IHR (see Methods for assumptions). Fitting the model to rounds 1-7 (1 May - 3 December 2020), we estimated the IHR to be 2.6% (2.5%, 2.7%), and the IFR to be 0.67% (0.65%, 0.70%). Fitting the model instead to rounds 14-19 (9 September 2021 - 1 April 2022), we estimated the IHR to be approximately 3.5-fold lower at 0.76% (0.75%, 0.77%), and the IFR to be approximately 7-fold lower at 0.097% (0.096%, 0.099%).

The severity of the virus, as measured per infectious individual, was found to be far lower in those aged 64 and under, relative to those aged 65 and over. From the models fitted to rounds 1-7 (1 May - 3 December 2020) we estimated the IHR to be 0.96% (0.93%, 1.00%) and the IFR to be 0.059% (0.057%, 0.061%) for those aged 64 and under. In comparison, the IHR and IFR for those aged 65 and over were approximately 16-fold and 91-fold higher, at 15% (14%, 17%) and 5.4% (4.9%, 5.9%) respectively for the same time period. As before, estimates of the IHR and IFR were found to be lower when the model was fitted to rounds 14-19 instead.

### Changes in severity during mass vaccination and the emergence of Alpha, Delta

We detected an increase in the severity of the virus into the winter of 2020. Using the best-fit time-lag obtained from fitting the time-delay model to rounds 1-7 (1 May - 3 December 2020) of REACT-1, we calculated the daily IFR and IHR from the modelled estimates of swab-positivity and the time-lag adjusted signals of hospitalisations and deaths over rounds 1-13 (1 May 2020 - 12 July 2021) of REACT-1 (Figure 6). An increase in the daily IFR was observed in late-November, with the increase lasting until late-January. We compared the mean IFR/IHR over approximately 4-week periods to a baseline period from 1 May 2020 to 11 November 2020 (Supplementary Table 3), a period before vaccinations or high proportions of any variants (Alpha, Delta, Omicron). The mean IFR in late-November 2020 (8 November – 5 December) was 1.68 (1.39, 1.93) times greater than baseline, and in January 2021 (3 January – 30 January) was 1.31 (1.11, 1.56) times greater than baseline; during these two periods the Alpha variant was responsible for 15%, and 86% of infections respectively. The increase in the IFR was observed in all age-groups, but no increase was observed in the IHR.

**Figure 6.**
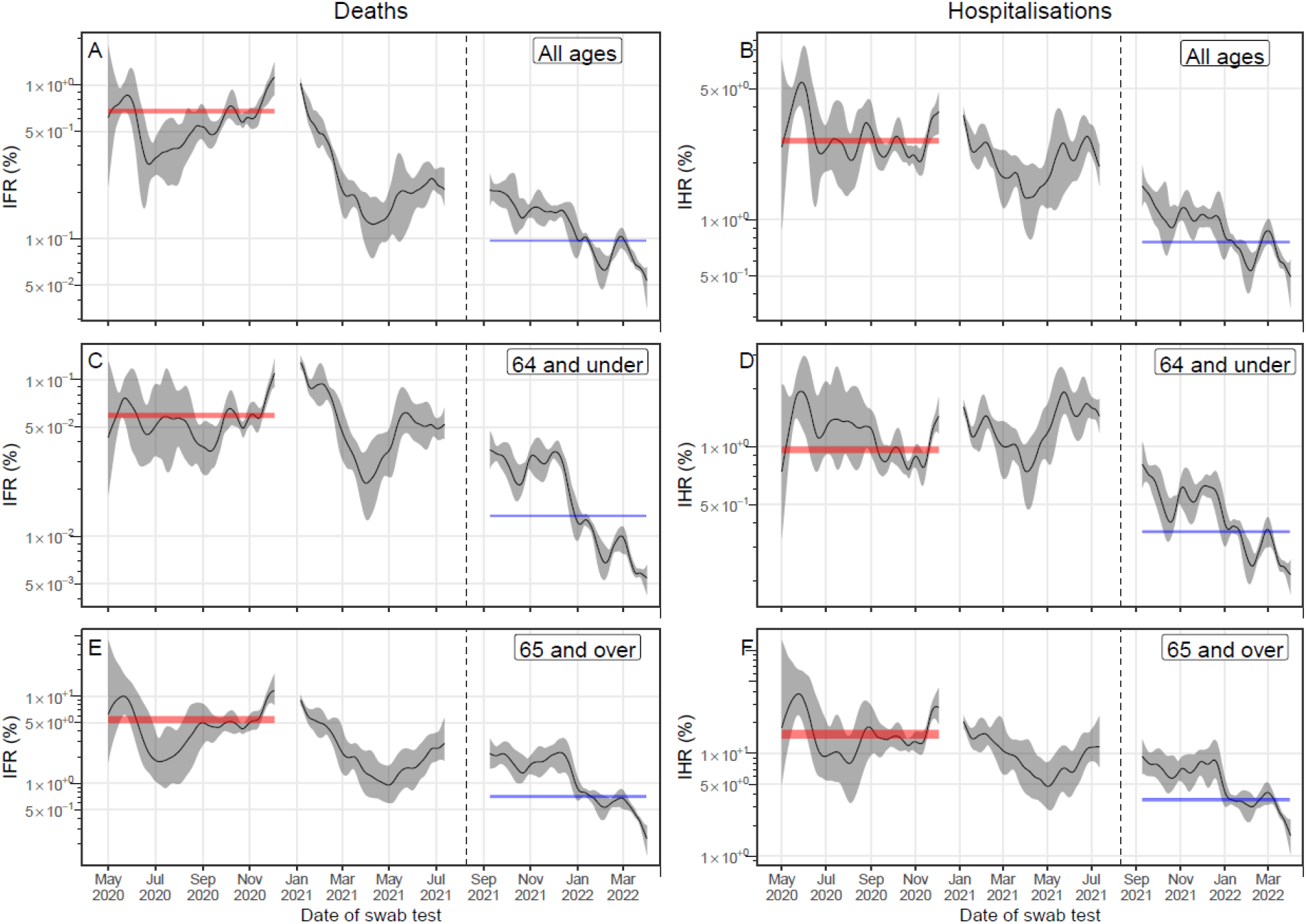
Estimates of the IFR and IHR over 19 rounds of REACT-1. Inferred IFR and IHR (solid black line, grey shaded region is 95\% credible interval) directly calculated from the multiplicative difference between the REACT-1 P-splines for swab positivity and the time-lag adjusted death/hospitalisations P-splines, accounting for population size, mean duration of positivity and test sensitivity. The 95% credible intervals of the best-fitting average IFR and IHR over rounds 1-7 (red shaded area) and rounds 14-19 (blue shaded area) estimated using time-delay models are shown for comparison. The black vertical dashed line on 10 August 2021 splits the data into two periods: rounds 1-13 and rounds 14-19 of REACT-1. (A) IFR across all age groups assuming a time-lag of 26 days during rounds 1-13 and 18 days during rounds 14-19. (B) IHR across all age groups assuming a time-lag of 19 days during rounds 1-13 and 7 days during rounds 14-19. (C) IFR in those aged 64 and under assuming a time-lag of 24 days during rounds 1-13 and 16 days during rounds 14-19. (D) IHR in those aged 64 and under assuming a time-lag of 17 days during rounds 1-13 and 6 days during rounds 14-19. (E) IFR in those aged 65 and over assuming a time-lag of 24 days during rounds 1-13 and 19 days during rounds 14-19. (F) IHR in those aged 65 and over assuming a time-lag of 18 days during rounds 1-13 and 9 days during rounds 14-19.

The severity of the virus, as measured by the IFR and IHR, decreased from late-January 2021 until April 2021, after which it increased until 12 July 2021 (the end of round 13). Both the daily IFR and daily IHR decreased until April. The mean IFR in April 2021 (28 March – 24 April) was significantly reduced to 0.25 (0.17, 0.34) of baseline, and the mean IHR to 0.51 (0.35, 0.68) of baseline. During this period of time the Alpha variant represented 96% of infections, 47% of the population had received at least one dose of vaccine, and 11% of the population had received two doses of the vaccine. By June/July 2021 (20 June – 17 July) the mean IFR had increased to 0.43 (0.37, 0.53) of baseline, and the mean IHR had increased to 0.84 (0.72, 1.03) of baseline. At this point in time the proportion vaccinated had increased to 66% with at least one dose and 50% with two doses; the Delta variant was now the dominant variant representing 99% of infections over this period.

There was a more substantial and quicker decrease in severity for those aged 65 and over, compared to those aged 64 and under. In April 2021 (28 March – 24 April), for those aged 65 and over the mean IFR and IHR were 0.28 (0.18, 0.49) and 0.42 (0.27, 0.74) of baseline respectively. For those aged 64 and under the mean IFR and IHR were comparatively higher at 0.48 (0.33, 0.71) and 0.72 (0.52, 0.97) of their baseline respectively. By June/July 2021 (20 June – 17 July) the mean IFR had increased to 0.98 (0.80, 1.15) of baseline for those aged 64 and under, but only to 0.63 (0.44, 1.06) for those aged 65 and over. The mean IHR in June/July 2021 had only increased to 0.76 (0.53, 1.24) for those aged 65 and over, but it was now significantly higher than baseline in those aged 64 and under, at 1.32 (1.14, 1.55) times baseline. There were large differences in the proportion vaccinated by this period; 98% of those aged 65 and over had received two doses of vaccine, whereas only 39% of those aged 64 and under had received two doses of vaccine.

### Changes in severity during booster vaccination and the emergence of Omicron

From September 2021 to April 2022 the severity of the virus decreased significantly. Using the best-fit time-lag obtained from fitting the time-delay model to rounds 14-19 of REACT-1, we calculated the daily IFR and IHR from the modelled estimates of swab-positivity and the time-lag adjusted signals of hospitalisations and deaths for rounds 14-19 of REACT-1 (Figure 6). The daily IFR and IHR decreased steadily from September 2021 onwards with a sharp and rapid reduction in late-December 2021. Over this period of time the proportion of the population that had received a third dose of vaccine (‘booster’ dose) steadily increased, saturating at about 54% by mid-January 2021 (Supplementary Table 4). Observed trends in the daily IFR and IHR were broadly similar across both age groups, despite approximately double the proportion of the population aged 65 and over having received a booster dose, compared to the population aged 64 and under, by the end of the study. The rapid reduction of the daily scaling parameters in late-December 2021 coincided with the rapid increase in the proportion of infections caused by the Omicron variant. We compared the mean IFR/IHR over approximately 4-week periods to a baseline period from 4 September 2021 to 16 October 2021 (Supplementary Table 4), a period before Omicron’s emergence and with low proportions of the population having received booster doses. By March 2022 (6 March – 2 April), when Omicron had reached near total coverage (responsible for 99% of infections), the mean IFR was 0.069% (0.066%, 0.072%) at 0.36 (0.31, 0.42) of baseline and the mean IHR was 0.62% (0.58%, 0.65%) at 0.50 (0.44, 0.59) of baseline.

## Discussion

This study demonstrates the clear temporal relationship between prevalence in the community, as measured by swab positivity, and severe outcomes. Due to the lag between the time series it suggests that large community testing studies such as REACT-1 could in the future be used not just for estimating prevalence but also in the short-term forecasting of severe outcomes. Previous analysis has suggested the time-delay between symptom onset and hospitalisations to be approximately 8 days [19,32,33] and between symptom onset and death to be approximately 16 days [18,19]; the time-delay found between REACT-1 swab positivity and these severe outcomes were both higher than these estimates during rounds 1-7 (May 2020 - December 2020) of the study. This might be due to REACT-1 better capturing asymptomatic and presymptomatic infections due to its sampling procedure.

During rounds 14-19 (September 2021 - April 2022) the time-delay between REACT-1 swab positivity and severe outcomes was shorter and more in line with the delays from symptom onset. This could suggest a change in the inherent biology of the virus, due to the newly emerged Omicron variant or due to the substantial buildup of immunity (due to vaccination and natural infection). It is also possible that the extremely high levels of prevalence observed in England over this period [17] caused a significant number of individuals that were hospitalised or went on to die of other causes to also be infected and test positive for SARS-CoV-2: the time series of severe outcomes would now be made up of two components, the true signal (due to infection leading to a severe outcome) and a pseudo-signal equivalent to the time series of prevalence. This would have the effect of reducing the time-lag, and may also explain the slight lower levels of correlation observed during this period.

In January 2021 the government’s mass vaccination campaign was substantially accelerated. Vaccines have been shown to be highly effective in preventing deaths related to COVID-19 [8] and so it is unsurprising that a few weeks after this the rate of deaths began to diverge from prevalence. This ‘decoupling’ was earlier and more significant in those aged 65 and over, most likely reflecting the vaccination campaign prioritising doses being given to the oldest individuals first. Though some degree of ‘decoupling’ was seen in the hospitalisation data it was less significant and occurred later. A potential explanation is that the vaccine has led to an overall reduction in the ‘severity pyramid’ of the virus and so those who might have died if unvaccinated were instead only hospitalised, delaying the reduction in hospitalisations. Another potential explanation is that the early vaccination of healthcare workers could have led to a reduction in nosocomial transmission leading to less infections in vulnerable patients and fewer deaths.

By measuring the difference between smoothed estimates of swab-positivity, and the time-lag adjusted signals for deaths and hospitalisations we were able to estimate the IFR and IHR of the SARS-Cov-2 virus over time. Though these estimates relied on some assumptions (see Methods), they showed strong agreement with other available estimates of the IFR and IHR over the period of 2020. We estimated an IHR of 2.6% during rounds 1-7 of REACT-1, this compared to an estimated IHR at the start of England’s first wave of 2.55% [23]. Our estimate of IFR over rounds 1-7 at 0.67% was slightly lower than the estimated IFR at the start of England’s first wave (1.00%), but was consistent with estimates at the end of England’s first wave which had the IFR at 0.79% (0.63%, 0.99%) [23].

There are large amounts of noise and correlation in the continuous estimates of the IFR and IHR making it difficult to assess overall trends. However, there does appear to have been a reduction in the IFR over the summer of 2020 matching some previous analysis [23]. There also appeared to be an upwards trend in the IFR and IHR into January 2021, this increase in lethality has been identified in other work [34] and is potentially driven by the increased severity of the Alpha variant [5], seasonality [35], or increased pressure on health services. However, due to the REACT-1 study not being in the field during December 2020 it is poorly placed to identify this as an estimate for the December peak in prevalence could not be made. Overall hospitalisations appeared to more closely follow prevalence than did deaths, even ignoring the divergences in 2021. This suggests a non-linearity between hospitalisations and deaths that could be a product of patterns in transmission, changing criteria for admissions or propensity to seek care.

The rapid increase in both the IFR and IHR from April 2021 likely reflects the emergence of the Delta variant and its rapid spread during a period with few new vaccinations among the most at-risk groups [11]. Vaccine effectiveness against Delta has been found to be reduced relative to previously circulating lineages after only a single dose, but broadly similar in those that have been double vaccinated [13]. This could explain the differences between age groups, with the IFR and IHR increasing to a far greater extent in those aged 64 and under, as by the end of April there was near total coverage of those aged over 65 with two doses of the vaccine. By July 2021 Delta accounted for almost all infections [12] and vaccine coverage in those aged over 65 was approximately constant. At this point in time the IFR for those aged over 65 was still higher than its lowest estimate in April suggesting that the Delta variant has also an increased severity over Alpha and other previously circulating lineages. It has previously been estimated that there is a ∼2 fold greater hazard of hospitalisation when infected with the Delta variant relative to Alpha [36,37], which could explain the large increase in the IHR even with high vaccine coverage.

A combination of booster vaccine doses and the emergence of the Omicron variant likely contributed to the reduction in both the IFR and IHR from September 2021 to April 2022. Booster doses have been found to be highly effective at reducing the odds of death or hospitalisation [38,39]. The exact contribution these made to reducing the IHR and IFR is unknown, though there appeared to be a mild reduction over the period booster doses were administered. Without booster doses there likely would have been an increase due to waning of vaccine effectiveness [25,38]. Omicron infections have been found to be less severe than Delta infections [40], which potentially explains the rapid reduction of the IFR and IHR in December (when Omicron rapidly replaced Delta as the dominant variant).

This study has limitations. A large duration of REACT-1 data was required to get an accurate estimate in the time-lag parameter, but this means changes in the time-lag over shorter periods can not be reliably detected. There is a possibility that vaccines and variants have had not only an effect on the severity of the virus, but also on the time-delay until a severe outcome in a way we have not accounted for in the analysis. Additionally, REACT-1 tests for swab-positivity and so is not actually a measure of when people are infected. At low prevalence there can be biases due to long term shedders [41] inflating the swab positivity measurements despite lower levels of infection than suggested. REACT-1 provides an accurate picture of prevalence in the community, but severe outcomes are more closely linked to prevalence in at risk individuals. Trends in prevalence in care homes during the pandemic have had some marked differences with community prevalence [23] which may have introduced biases. Though our model assumes a straightforward time-delay between prevalence and severe outcomes (a convolution with a delta function) it is more likely a transformation involving a convolution with a more dispersed shape. This may lead to trends in prevalence over short time-scales being smoothed out in the resulting death and hospitalisation time series.

## Conclusion

Over the duration of the pandemic in England there has been a decline in the severity of the SARS-CoV-2 virus. However, there remain many populations worldwide that have lower vaccination rates, and where severity may not have reduced to as great an extent. We have observed that the severity of the virus can vary when new variants are introduced. Though the current Omicron variant seems to have led to a reduction in severity, there remains the distinct possibility that future variants will lead to an increase. In preparing for future waves of SARS-CoV-2 infection, it remains paramount that any increases in the severity of the virus are detected rapidly. Prevalence studies such as REACT-1 can provide unbiased estimates of infection levels in a population over time allowing any changes in the IFR or IHR to be identified quickly. With advanced warning, appropriate interventions can then be implemented when they are most effective, and additional vaccine doses can be administered to ensure the link between severe outcomes and prevalence is as small as possible into the future.

## Methods

### REACT-1 Data

The study protocol of REACT-1 has been well described [17,29]. Each round a random subset of the population, chosen at the lower tier local authority (LTLA, N=315) level using the list of GP patients in England held by the NHS, is sent a letter inviting them to participate. Those who agree to participate are then sent a swab-test which they self administer (for 5-12 year olds a parent/guardian will administer the test). The participants also complete a questionnaire providing socio-demographic information such as age, gender, ethnicity, etc.

Swab tests are collected by a courier and sent via cold chain to a private lab for a RT-PCR test to be performed. From the test we determine if someone is swab positive or not using a definition of either both N- and E-gene detected, or just N-gene detected, but with a Ct value less than 37.

### Public data

Data for deaths and hospitalisations is obtained from ‘the official UK government website for data and insights on coronavirus’ [6]. Daily hospitalisation figures include all people in England who are admitted to hospital within 14 days of testing positive for COVID-19, and those who test positive after admission. Daily death figures include the number of people in England that died within 28 days of their first positive test for COVID-19 being reported (by date of death not date of reporting). Daily death data can be updated as new information is provided, this means the most recent days are usually unreliable, we corrected for this by not including the last 5 days in the data downloaded. Both hospitalisation and death data were available by age group; From this we created three time series for each data set: the total number of events (death or hospitalisation), the number in those aged 64 and under, and the number in those aged 65 and over. Death and hospitalisation data were downloaded on 20 May 2022 and only data up to 15 May 2022 was included in the analysis.

The daily cumulative number of individuals that have been vaccinated with a single dose and with two doses (any vaccine approved in the UK), by age group was again downloaded from ‘the official UK government website for data and insights on coronavirus’ [6]. Data by age group was collated into those aged 64 and under, those aged 65 and over, and all age groups. The cumulative number of vaccinations given (first and second doses) was then converted into the cumulative proportion of the population vaccinated using population estimates of England by age group [42]. Note that current population estimates are unavailable and are most likely greater than the values used; estimates of the proportion of the population vaccinated are thus not exact, but still informative.

The weekly numbers of each SARS-CoV-2 lineage detected in routine surveillance data by lower tier local authority (LTLA) was downloaded from the ‘Lineage in Space and Time website’ [43]. Data by LTLA was aggregated in order to give the daily number of each lineage detected in England as a whole. The weekly proportion of lineages that were the Alpha variant (or an alpha sub-lineage), the Delta variant (or a Delta sub-lineage) and the Omicron variant (or an omicron sub-lineage) was then straightforwardly calculated.

### P-spline model + mixed-effects P-spline

Smoothed models were fit to each time series in order to get an estimate of the expected number of outcomes (deaths, hospitalisations) or the expected prevalence (REACT-1 swab positivity). Bayesian P-spline models [44,45] were fit to the hospitalisation and death data for each age group (all, 64 or less, 65 and over), and to the overall swab positivity for REACT-1. Due to the reduction in power in the REACT-1 data when splitting it into two age groups, a single mixed-effects Bayesian P-spline model was fit to the swab positivity in REACT-1 for the two age groups. The models consist of a system of basis splines defined over the window of the study period with approximately one basis spline every 5 days. The P-spline model is then a linear combination of these basis splines:

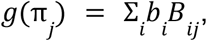

where π_*j*_ is the outcome variable of interest on the *j*^*th*^ day, *g*()is the link function (logit for binomial swab positivity data, log for count data), *b*_*i*_ is the coefficient for the *i*^*th*^ basis-spline and *B*_*ij*_ the value of the *i*^*th*^ basis spline on the *j*^*th*^ day. Overfitting of the model was prevented through the inclusion of a second-order random walk prior on the basis spline coefficients, *b*_*i*_:

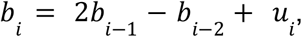

where

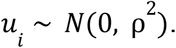

This prior penalises any changes in the first derivative of the response function, reflecting the expected trend of an epidemic over a small time period (constant growth rate). The degree to which changes in the first derivative are penalised is controlled by ρ which is a further parameter of the model and takes a loose but proper inverse gamma prior ρ ∼ *IG*(0. 001, 0. 001). The first two basis spline coefficients are given an uninformative prior, *b*_1,_ *b*_2_ ∼ *Constant*.

In the mixed-effects version of the model in which the model fits to the time series for two age-groups simultaneously the second-order random-walk prior is modified slightly:

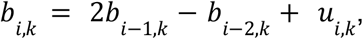

where

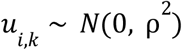

and

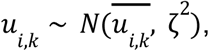

Where 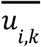 is the mean value of *u*_*i,k*_ averaged over *k* age groups. For the most part this the same as before but now parameters are defined for *k* age-groups. The difference lies in that as well as penalising changes in the first derivative of the response function, differences between age groups in the changes of the first derivative are also penalised. This has the effect of syncing the model across age-groups whilst also allowing divergences to occur when there is sufficient evidence. The degree of penalisation is controlled by a further parameter *ζ* which we give a loose but proper inverse-gamma prior as before, *ζ* ∼ *IG*(0. 001, 0. 001).

All models are fit to data using a No-U-Turn Sampler (NUTS) [46] implemented in STAN [47]. For the REACT-1 data we fit the daily weighted number of positive and negative tests assuming a Binomial likelihood. When fitting to the daily number of deaths and hospitalisations we assume a Negative-Binomial likelihood with an overdispersion parameter that is treated as an additional parameter of the model with an uninformative constant prior. The model fitting returns a full posterior distribution of all parameters from which the mean response function and credible intervals can be calculated. Estimates of swab positivity for the period between REACT-1 rounds 7 and 8 are not included in any analysis as between the rounds there was a large peak in infections that we cannot estimate using the REACT-1 data. Similarly we did not include estimates of swab positivity for the period between rounds 13 and 14 in any analysis; this is as there was a substantial break in the study (∼2 months) for which dynamics cannot be inferred accurately. Estimates of swab positivity between other rounds are included as there was only a small break between most rounds (∼2 weeks) that the P-spline model can effectively infer over.

### Time-delay model

In order to investigate the relationship between the REACT-1 data and hospitalisations and deaths we define a simple model consisting of two parameters. The first parameter, the time-lag τ sets the discrete number of days between the REACT-1 data and the time series of interest. The second parameter the scaling factor ε sets the multiplicative difference between REACT-1 data and the time series of interest correcting for population size. The estimated proportion swab positive on day *i* is then related to the time series of interest on day *i* + τ by:

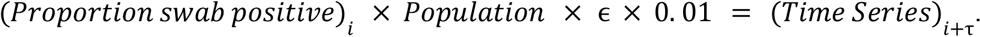

Written this way the scaling parameter, ε, represents the percentage of those in the population that are swab-positive on day *i* that will be admitted to hospital or dead on day *i* + τ. The modelled proportion swab-positive on day *i*:

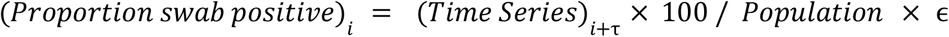

can then be fitted to the REACT-1 daily weighted number of positive and negative tests assuming a Weighted-Binomial likelihood. If this was done using the raw data for hospitalisations or deaths then there would be errors for any days in which zero counts occurred. There would also be the possibility for overfitting due to well-aligned noise between the raw time series and the REACT-1 data. In order to avoid these pitfalls, instead of using the raw data as the time series, the P-spline model estimates are used instead. To take into account the uncertainty in the P-spline model, a random subset of 1000 parameter combinations is selected from the posterior distribution. For each set of parameter combinations a time-series can be calculated. The average log-likelihood, fitting to the REACT-1 data, over all 1000 random draws of the P-spline’s posterior is then used to fit the time-lag and scaling parameter using an MCMC. The time-delay models were fit to all rounds of REACT-1 and to subsets of rounds: rounds 1-7, rounds 8-13, rounds 14-19. Further analyses were only performed using the time-lags from models fit to rounds 1-7 and rounds 14-19.

### Correlation between time series

Under the assumption of a particular time-lag, the Pearson correlation was measured between the mean daily swab positivity (as estimated using the P-spline model) and the time-delayed mean daily severe outcomes (deaths or hospitalisations) (as estimated using the P-spline model). Correlation between swab positivity and severe outcomes was estimated for the periods of rounds 1-7, rounds 8-13 and rounds 14-19. The time-lags used were those estimated from the time-delay model fit to rounds 1-7 and fit to rounds 14-19.

### Variation in scaling parameter over time

With the assumption of a particular time-lag, and that it does not change over time, variation in the scaling parameter over time between REACT-1 data and hospitalisation/death data can straightforwardly be estimated. For each day the posterior distribution for the estimate of swab-positivity that day, inferred from the P-spline model fit to REACT-1, can be extracted. Similarly the posterior distribution for the estimate of hospitalisations/deaths on a fixed number of days later (determined by time-lag) can also be extracted. The daily scaling parameter, and its uncertainty, between the two P-spline estimates can then be calculated for the whole study period.

The average scaling parameter over a set period of time can also readily be calculated. We split the time period of rounds 1-13 of REACT-1 into approximately 4-week periods and one initial baseline period running from 1 May 2020 - 7 November 2020. This period was chosen as the baseline as it occurred before any vaccination had occurred and before the Alpha variant had increased to a substantial proportion. Similarly we split the time period of rounds 14-19 of REACT-1 into approximately 4-week periods and one initial baseline period running from 4 September 2021 - 16 October 2021. This period was chosen as the baseline as it occurred before Omicron emerged and before the proportion receiving a third vaccine dose was substantial.

For each period, the posterior distribution for the estimates of swab positivity over the period, inferred from the P-spline model fit to REACT-1, can be extracted. The posterior distribution of the mean swab positivity over the period can then simply be calculated. Similarly the posterior distribution for the estimate of hospitalisations/deaths for the equivalent period a fixed number of days later (determined by time-lag) can also be extracted and the posterior of the mean calculated. The mean scaling parameter (the difference between mean swab positivity and mean hospitalisations/deaths) over the period, and its uncertainty, can then be calculated. Additionally the multiplicative difference between the mean scaling parameter for a specific period and the baseline period, and its uncertainty can be calculated.

### Converting the scaling parameters to the IFR/IHR

The scaling parameters, defined as they are, correspond to the percentage of those who are swab-positive in a population on a particular day that will be hospitalised/dead on a day in the future set by the time-lag parameter. If the swab-positivity can be converted to the incidence of infection, then the scaling parameter can be converted to the IFR/IHR. The time-series of incidence and swab-positivity have been shown to be remarkably similar - though with a time-delay due to the finite duration individuals remain swab-positive for [48]. Under the simplifying assumption that swab-positivity can be converted to incidence by a multiplicative constant, the IFR and IHR can similarly be estimated by multiplying the scaling parameter by the same constant. The constant we use consists of the mean duration an individual remains positive for, estimated at 14.0 days, and the sensitivity of the REACT-1 study, estimated at 0.79 [48]. These two numbers allow all scaling parameter estimates to be converted to IFR (for deaths) and IHR (for hospitalisations) by multiplying by 11.06.

## Data Availability

Access to individual level REACT-1 data is restricted due to ethical and security considerations. Summary statistics and data, including the weighted daily number of positive tests and daily total number of tests, are available at https://github.com/mrc-ide/reactidd/tree/master/inst/extdata. Additional summary statistics and results from the REACT-1 programme are also available at https://www.imperial.ac.uk/medicine/research-and-impact/groups/react-study/real-time-assessment-of-community-transmission-findings/. REACT-1 study materials are available for each round at https://www.imperial.ac.uk/medicine/research-and-impact/groups/react-study/react-1-study-materials/.

https://github.com/mrc-ide/reactidd/tree/master/inst/extdata

https://www.imperial.ac.uk/medicine/research-and-impact/groups/react-study/real-time-assessment-of-community-transmission-findings/

https://www.imperial.ac.uk/medicine/research-and-impact/groups/react-study/react-1-study-materials/

## Acknowledgements

SR, CAD acknowledge support: Medical Research Council (MRC) Centre for Global Infectious Disease Analysis, National Institute for Health Research (NIHR) Health Protection Research Unit (HPRU), Wellcome Trust (200861/Z/16/Z, 200187/Z/15/Z), and Centres for Disease Control and Prevention (US, U01CK0005-01-02). GC is supported by an NIHR Professorship. HW acknowledges support from an NIHR Senior Investigator Award and the Wellcome Trust (205456/Z/16/Z). PE is Director of the MRC Centre for Environment and Health (MR/L01341X/1, MR/S019669/1). PE acknowledges support from Health Data Research UK (HDR UK); the NIHR Imperial Biomedical Research Centre; NIHR HPRUs in Chemical and Radiation Threats and Hazards, and Environmental Exposures and Health; the British Heart Foundation Centre for Research Excellence at Imperial College London (RE/18/4/34215); and the UK Dementia Research Institute at Imperial (MC_PC_17114). We thank The Huo Family Foundation for their support of our work on COVID-19. MC-H acknowledge support from Cancer Research UK, Population Research Committee Project grant ‘Mechanomics’ (grant No 22184 to MC-H). MC-H acknowledges support from the H2020-EXPANSE (Horizon 2020 grant No 874627) and H2020-LongITools (Horizon 2020 grant No 874739).

We thank key collaborators on this work – Ipsos MORI: Kelly Beaver, Sam Clemens, GaryWelch, Nicholas Gilby, Kelly Ward and Kevin Pickering; Institute of Global Health Innovationat Imperial College: Gianluca Fontana, Didi Thompson and Lenny Naar; Molecular Diagnostic Unit, Imperial College London: Prof. Graham Taylor; Patient Experience Research Centre at Imperial College and the REACT Public Advisory Panel; NHS Digital for access to the NHS register; and the Department of Health and Social Care for logistic support.

## Ethics

The REACT-1 study received research ethics approval from the South Central-Berkshire BResearch Ethics Committee (IRAS ID: 283787).

## Data availability

Computer code supporting the paper is available at https://github.com/mrc-ide/reactidd and also on zenodo (DOI:10.5281/zenodo.7085123)

## Supplementary Tables

All supplementary tables are available in the supplementary materials. Legends for the supplementary tables are provided below.

**Supplementary Table 1**. Estimated time-lags and IFR/IHR for all time-delay models fit to REACT-1.

**Supplementary Table 2**. Pearson correlation between modelled estimates of swab positivity and modelled estimates of deaths and hospitalisations.

**Supplementary Table 3**. The average proportion of population vaccinated, average proportion of infections caused by each variant, and mean IFR and IHR over fixed periods of time over the duration of rounds 1-13 of REACT-1. Mean IFR/IHR for each period are compared to a baseline period running from 1 May 2020 - 7 November 2020. The time-lags between positivity and deaths/hospitalisations used in calculating mean IFR and IHR are the best-fit time-lags from the time-delay models fit to rounds 1-7 of REACT-1.

**Supplementary Table 4**. The average proportion of the population vaccinated, average proportion of infections caused by each variant, and mean IFR and IHR over fixed periods of time over the duration of rounds 14-19 of REACT-1. Mean IFR/IHR for each period are compared to a baseline period running from 4 September 2021 - 16 October 2021. The time-lags between positivity and deaths/hospitalisations used in calculating mean IFR and IHR are the best-fit time-lags from the time-delay models fit to rounds 14-19 of REACT-1.

